# Comparisons of Heart Rate Variability Responses to Head-up Tilt With and Without Abdominal and Lower-Extremity Compression in Healthy Young Individuals: A Randomized Crossover Study

**DOI:** 10.1101/2023.07.24.23293087

**Authors:** Kazuaki Oyake, Miyuki Katai, Anzu Yoneyama, Hazuki Ikegawa, Shigeru Kani, Kimito Momose

## Abstract

**Introduction:** Abdominal and lower-extremity compression techniques can help reduce orthostatic heart rate increase. However, the effects of body compression on the cardiac autonomic systems, which control heart rate, remain unclear. This study aimed to compare heart rate variability, a reflection of cardiac autonomic regulation, during a head-up tilt test with and without abdominal and lower-extremity compression in healthy young individuals.

**Methods:** In a randomized crossover design, 39 healthy volunteers (20 females, aged 20.9 ± 1.2 years) underwent two head-up tilt tests with and without abdominal and lower-extremity compression. Heart rate and heart rate variability parameters were measured during the head-up tilt tests, including Stress Index, root mean square of successive differences between adjacent R-R intervals, low- and high-frequency components, and low-to-high frequency ratio.

**Results:** Abdominal and lower-extremity compression reduced the orthostatic increase in heart rate (p < 0.001). The tilt-induced changes in heart rate variability parameters, except for the low-frequency component, were smaller in the compression condition than in the no-compression condition (p < 0.001). Additionally, multiple regression analysis with potentially confounding variables revealed that the compression-induced decrease in Stress Index during the head-up tilt position was a significant independent variable for the compression-induced reduction in heart rate in the head-up tilt position (coefficient = 0.411, p = 0.025). Comparative analyses revealed that abdominal and lower-extremity compression has a notable impact on the compensatory sympathetic activation and vagal withdrawal typically observed during orthostasis, resulting in a reduction of the increase in heart rate. Furthermore, this decrease in heart rate was primarily attributed to the attenuation of cardiac sympathetic activity associated with compression.

**Conclusion:** Our findings could contribute to the appropriate application of compression therapy for preventing orthostatic tachycardia. This study is registered with UMIN000045179.

## INTRODUCTION

Assuming an upright position increases gravitational forces and leads to pooling of 500–800 mL of blood in the venous system, pelvic and splanchnic circulation, and lower extremities, which reduces venous return, stroke volume, and mean blood pressure. The subsequent compensatory response includes sympathetic activation and vagal withdrawal, leading to increased heart rate, cardiac contractility, and peripheral vascular resistance (Mosqueda-Garcia et al., 2000).

External compression applied to the abdomen and lower extremities can aid in redirecting pooled blood back to the heart and potentially attenuate orthostatic increase in heart rate (Lee et al., 2018). Thus, compression garments have been used as a potential treatment for postural orthostatic tachycardia syndrome (POTS) (Heyer, 2014; Bourne et al., 2021). POTS is a chronic form of orthostatic intolerance predominantly affecting females of child-bearing age, which is characterized by frequent symptoms of dizziness and palpitations upon standing, accompanied by an increase in heart rate of 30 bpm or more, and the absence of orthostatic hypotension (Freeman et al., 2011; Sheldon et al., 2015; Arnold et al., 2018). In addition, POTS is often associated with symptoms of depression, anxiety, sleep disturbances, cognitive dysfunction, exercise intolerance, functional disability, and impaired health-related quality of life (Bagai et al., 2011; Shibata et al., 2012; Anderson et al., 2014; Pederson and Brook, 2017; Hutt et al., 2020; Rich et al., 2022; Vas et al., 2022). Nonpharmacologic therapies, including the use of compression garments, are often recommended as a first-line treatment for POTS (Bryarly et al., 2019). However, to the best of our knowledge, the effects of compression on the cardiac autonomic systems controlling heart rate have not been investigated, even in healthy individuals. From an ethical and scientific perspective, it is crucial to first understand the cardiac autonomic responses associated with the reduction in orthostatic heart rate increase with compression in individuals without POTS.

Analysis of heart rate variability is widely used as a standard method for assessing cardiac autonomic nervous functions (Camm et al., 1996; Laborde et al., 2017; Shaffer and Ginsberg, 2017). Therefore, this study aimed to compare heart rate variability responses to head-up tilt (HUT) with and without abdominal and lower-extremity compression in healthy young individuals. A systematic review has highlighted a lower root mean square of successive differences between adjacent R-R intervals (RMSSD), which is the primary time-domain measure of cardiac vagal tone, during the HUT position in individuals with POTS than healthy individuals (Swai et al., 2019). Based on existing knowledge, we hypothesized that abdominal and lower-extremity compression would effectively reduce the orthostatic decrease in RMSSD and the subsequent increase in heart rate compared to no compression. These findings would have substantial implications for the appropriate use of compression therapy in preventing the onset of orthostatic tachycardia.

## MATERIALS AND METHODS

### Study design

This study used a randomized crossover design. Participants underwent two consecutive HUT tests under no-compression and compression conditions. A 10-min supine rest period was designated between each test as a washout period. Participants were randomly assigned to either the no-compression or compression condition in the first trial using computer-generated random numbers. The study protocol was approved by the appropriate ethics committee of Shinshu University (approval number: 5254). All participants provided written informed consent before enrolment in the study. The study was performed per the 1964 Declaration of Helsinki, as revised in 2013.

### Participants

Recruitment was conducted by placing posters in the university campus. The inclusion criteria comprised age 20–40 years, no underlying disease, no history of syncope, and no smoking habit. The exclusion criteria were as follows: limited range of motion and/or pain that affects the HUT test, having any contraindications of medical compression treatment (Rabe et al., 2020), and taking any medication that interferes with cardiovascular control.

### Abdominal and lower-extremity compression

In the compression condition, participants were outfitted with an inflatable abdominal band along with medical compression stockings. The inflatable abdominal band consisted of an outer band made of hard polyester cloth with Velcro straps and an inner inflatable cuff of an aneroid sphygmomanometer (Tanaka et al., 1997). The band was attached to participants around the lower abdomen between the pubis and umbilicus (Smit et al., 2004; Figueroa et al., 2015) and inflated to 35–45 mmHg to compress the abdominal wall. The medical compression stockings used in this study (Jobst Bellavar class 3; BSN-JOBST GmbH, Emmerich am Rhein, Germany) delivered approximately 35–45 mmHg of pressure at the ankle. Appropriately sized stockings were selected according to the circumference of the ankle and calf, as recommended by the manufacturer.

### HUT test

Two trained assessors performed the HUT tests in a quiet room at a comfortable temperature. Participants were asked to refrain from eating and consuming caffeinated products for at least 2 h and to avoid vigorous physical activity for at least 12 h before each test (Cheshire and Goldstein, 2019; Finucane et al., 2019). The tests were performed between 5:00 and 7:00 p.m.

In the HUT test, participants remained in a resting supine position on a motorized tilt table (TB-653; Takada Ned Manufacturing Co., Ltd., Osaka, Japan) for 5 min before the postural change. After the 5-min supine rest, the tilt table was elevated to an angle of 70° for approximately 30 s and was maintained for 10 min (Cheshire and Goldstein, 2019; Bourne et al., 2021). If a participant experienced a severe symptom, such as presyncope, the test was promptly halted, and the participant was repositioned to a supine position. Throughout the HUT tests, participants were instructed to regulate their breathing at a rate of 0.25 Hz (approximately 15 breaths per minute) using a computer metronome. This measure was implemented to minimize any potential influence of respiratory fluctuations on heart rate variability assessments (Schipke et al., 1999; Laborde et al., 2017). The audio cue was available to help participants pace their breathing.

### Measurements and data analysis

The R-R intervals, the time interval between two consecutive R waves in the electrocardiogram, were continuously recorded during the HUT test using a portable 3-lead electrocardiograph monitor (Active Tracer AC-301A; GMS Inc., Tokyo, Japan) with a sampling rate of 1 kHz. Blood pressure was measured every minute on the left arm using an automated sphygmomanometer (HEM-907; Omron Co., Ltd., Kyoto, Japan). At the end of each test, participants were instructed to report the severity of orthostatic symptoms, such as dizziness and palpitations, that occurred during 10 min of HUT on a visual analog scale. The possible score ranged from 0 (no symptoms) to 100 (syncope or presyncope), measured in millimeters on a 100-mm horizontal line using a pen.

Heart rate and heart rate variability parameters were derived from the R-R intervals recorded during the last 5 min of the supine and HUT phases. These calculations were performed using Kubios HRV standard software version 3.5 (Kubios Oy, Kuopio, Finland). RMSSD was analyzed as the time-domain measure of heart rate variability to estimate vagal activity (Camm et al., 1996; Laborde et al., 2017; Shaffer and Ginsberg, 2017). Furthermore, the Stress Index, which is the square root of Baevsky’s stress index (Baevsky and Chernikova, 2017), served as an indicator of sympathetic activity (Ali et al., 2021; Software, 2021). Baevsky’s stress index was calculated using the following formula:

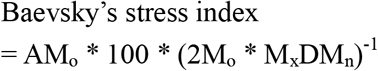

where the mode (M_o_) is the most frequent R-R interval expressed in seconds. The amplitude of the mode (AM_o_) was determined by employing a 50-ms bin width, quantified as the percentage of R-R intervals within the bin containing the mode in relation to the total number of R-R intervals recorded. Variation range (M_x_DM_n_) represents the disparity between the longest (M_x_) and shortest (M_n_) values of R-R intervals, measured in seconds. Enhanced sympathetic activation of the heart leads to a more stable heart rhythm, resulting in reduced variability in R-R intervals and an increased occurrence of R-R intervals with similar durations. Thus, when cardiac sympathetic activity increases, the histogram of R-R intervals becomes narrower and increases in height, leading to a higher Stress Index. Additionally, as power spectral analysis of the R-R intervals has also been widely used to quantify cardiac autonomic regulation (Camm et al., 1996; Laborde et al., 2017; Shaffer and Ginsberg, 2017), the frequency-domain parameters of heart rate variability were calculated using the fast Fourier transform model. Fast Fourier transform analysis used a Welch periodogram method with a window width of 256 seconds and 50% window overlap. Low-frequency (0.04–0.15 Hz) and high-frequency (0.15–0.40 Hz) spectral components were collected in absolute values of power (ms^2^). The low-frequency heart rate variability (LF) component reflects a complex and not easily discernible mix of sympathetic, parasympathetic, and other unidentified factors (Billman, 2013). The high-frequency heart rate variability (HF) component reflects the vagal tone and is more affected by respiratory rate than RMSSD (Laborde et al., 2017; Shaffer and Ginsberg, 2017). Furthermore, the LF/HF ratio was calculated from the absolute values of LF of HF. The LF/HF ratio is often regarded as an indicator of the sympathetic-parasympathetic balance, despite receiving some criticism for this interpretation (Billman, 2013).

The averaged values of blood pressure variables during the supine and HUT periods were also calculated from data collected during the final 5 min of each period. The changes in hemodynamic and heart rate variability parameters from supine to HUT were calculated by subtracting the values measured during the supine position from those obtained during the HUT position.

### Statistical analysis

The G Power computer program version 3.1.9.2 (Heinrich Heine University, Dusseldorf, Germany) (Erdfelder et al., 1996) was used for sample size calculation to detect the differences in hemodynamic and heart rate variability responses to HUT between the with and without-compression conditions. Based on a previous study that compared hemodynamic responses to HUT with and without lower-extremity compression in healthy adults (Lee et al., 2018), we used an estimated effect size of 0.80 for the paired t-test. Considering a statistical power of 0.80, an alpha level of 0.05, and an effect size of 0.80, the estimated sample size required was 15. Previous studies investigating the effectiveness of compression in reducing orthostatic tachycardia in individuals with POTS have primarily included female participants, accounting for more than 90% of the sample (Heyer, 2014; Bourne et al., 2021). This gender imbalance has limited the generalizability of the findings to males. Thus, to facilitate subgroup analysis comparing heart rate and heart rate variability responses to HUT between the no-compression and compression conditions based on participant sex, a minimum of 15 males and 15 females was deemed necessary.

Hemodynamic and heart rate variability parameters were subjected to a two-way repeated-measures analysis of variance (ANOVA) with two positions (supine and HUT) and two conditions (no compression and compression) as within-subject factors. A paired t-test with Bonferroni correction was performed to compare hemodynamic and heart rate variability parameters between the conditions in each position. In the subgroup analyses by sex, the changes in heart rate and heart rate variability parameters from supine to HUT were compared with and without compression using a paired t-test.

To explore the heart rate variability parameters associated with the compression-induced reduction in standing heart rate, we calculated the difference in heart rates during the HUT with and without compression as the dependent variable. Regarding the heart rate variability parameters with a significant interaction between position and condition, we also calculated the differences in the measurements during the HUT position with and without compression as the independent variables. The correlations between the dependent and independent variables were determined using Pearson’s product-moment correlation coefficient. In addition, to identify potential confounding variables, we examined the associations of the compression-induced change in heart rate with age, height, weight, body mass index, and the changes in systolic and diastolic blood pressure during the HUT position with compression using Pearson’s product-moment correlation coefficient and unpaired t-test based on variable types. We performed multiple regression analysis with forced entry to confirm whether the associations between the changes in heart rate and heart rate variability parameters observed in the bivariate analysis remained significant, even when adjusting for potential confounding variables. Statistical analyses were performed using GraphPad Prism version 9.00 for Windows (GraphPad Software, San Diego, California, USA). Statistical significance was set at a p-value less than 0.05.

## RESULTS

### Participants

A total of 40 healthy volunteers participated in this study. Owing to an inadequate R-R interval signal, one participant was excluded from the analysis. Consequently, data concerning 39 participants were included in the analysis. Table 1 shows the participants’ characteristics. A flow chart of the participants enrolled in this study is shown in Figure 1. According to the randomized sequence, 18 participants underwent the compression condition before the no-compression condition, whereas 21 participants experienced the no-compression condition preceding the compression condition.

**Table 1.** Participants’ characteristics.

| Variable | Overall<br>(n = 39) | Male<br>(n = 19) | Female<br>(n = 20) |
| --- | --- | --- | --- |
| Age (years) | 20.9 ± 1.2 | 21.5 ± 1.4 | 20.5 ± 0.6 |
| Height (m) | 1.65 ± 0.08 | 1.71 ± 0.05 | 1.59 ± 0.05 |
| Weight (kg) | 55.6 ± 8.5 | 61.0 ± 6.9 | 50.5 ± 6.4 |
| Body mass index (kg/m<sup>2</sup>) | 20.4 ± 2.1 | 21.0 ± 2.0 | 19.9 ± 2.2 |

Values are presented as mean ± standard deviation.

**Figure 1.**
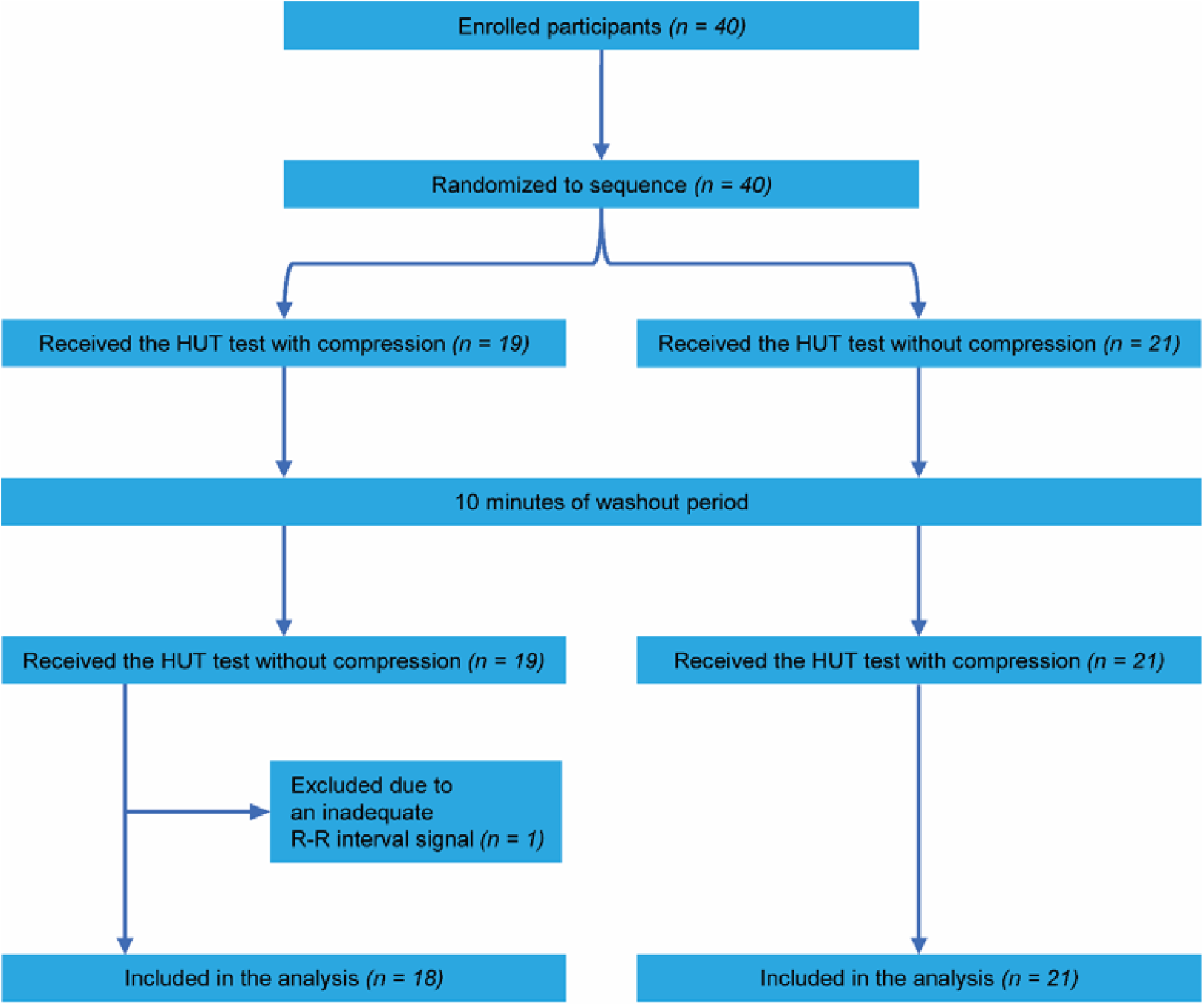
Flow diagram of participant enrolment HUT, head-up tilt

### Comparisons of hemodynamic and heart rate variability responses to HUT with and without abdominal and lower-extremity compression

All participants completed 10-min HUT in the no-compression and compression conditions without significant adverse events. The median (interquartile range) visual analog scale scores in the no-compression and compression conditions were 15 (5–41) mm and 10 (3–23) mm, respectively. Figure 2 illustrates hemodynamic and heart rate variability responses to HUT.

**Figure 2.**
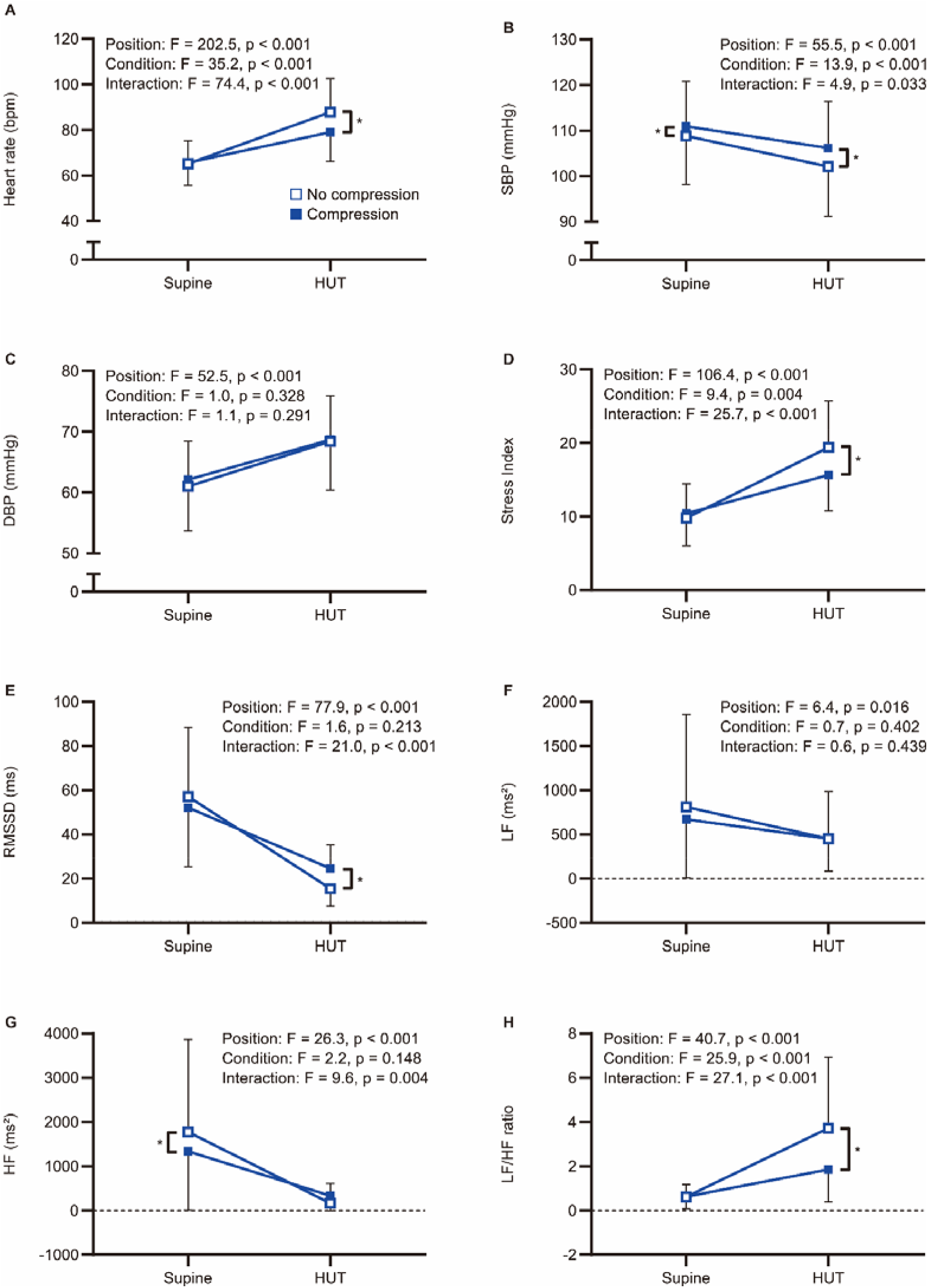
Comparisons of the hemodynamic and heart rate variability responses to head-up tilt with and without compression Hemodynamic and heart rate variability responses to head-up tilt including (A) heart rate, (B) systolic blood pressure, (C) diastolic blood pressure, (D) Stress Index, (E) RMSSD, (F) LF, (G) HF, and (H) LF/HF ratio. An asterisk indicates a significant difference between the conditions by Bonferroni multiple comparison test (p < 0.05). HUT, head-up tilt; SBP, systolic blood pressure; DBP, diastolic blood pressure; RMSSD, root mean square of successive differences between adjacent R-R intervals; LF, low-frequency component of heart rate variability; HF, high-frequency component of heart rate variability; LF/HF ratio, low-to-high frequency ratio

### Hemodynamic parameters

Representative data of heart rate changes during the HUT test in the no-compression and compression conditions are shown in Figure 3. The two-way repeated-measures ANOVA revealed significant main effects for position [F(1,38) = 202.5, p < 0.001] and condition [F(1,38) = 35.2, p < 0.001] on heart rate, as well as a significant interaction between position and condition [F(1,38) = 74.4, p < 0.001] (Figure 2A). Post-hoc pairwise comparisons revealed that there was no significant difference in supine heart rates between the conditions (t = 0.687, p = 0.993), while the heart rate during the HUT position was significantly lower in the compression condition than that in the no-compression condition (t = 11.510, p < 0.001). These results indicate that the increase in heart rate during the HUT test was significantly smaller in the compression condition than that in the no-compression condition. Six participants (five males and one female) showed a heart rate increase of ≥30 bpm from supine to HUT in the no-compression condition, while no participants exhibited an orthostatic heart rate increase of ≥30 bpm in the compression condition. Subgroup analyses revealed that the increase in heart rate during the test was significantly smaller in the compression condition than that in the no-compression condition, regardless of sex (p < 0.001) (Figure 4A). In addition, the supplementary analysis also indicated a significantly smaller heart rate increase from supine to HUT in the compression condition than that in the no-compression condition, regardless of whether the no-compression or compression condition occurred first (p < 0.001) (Figure 5).

**Figure 3.**
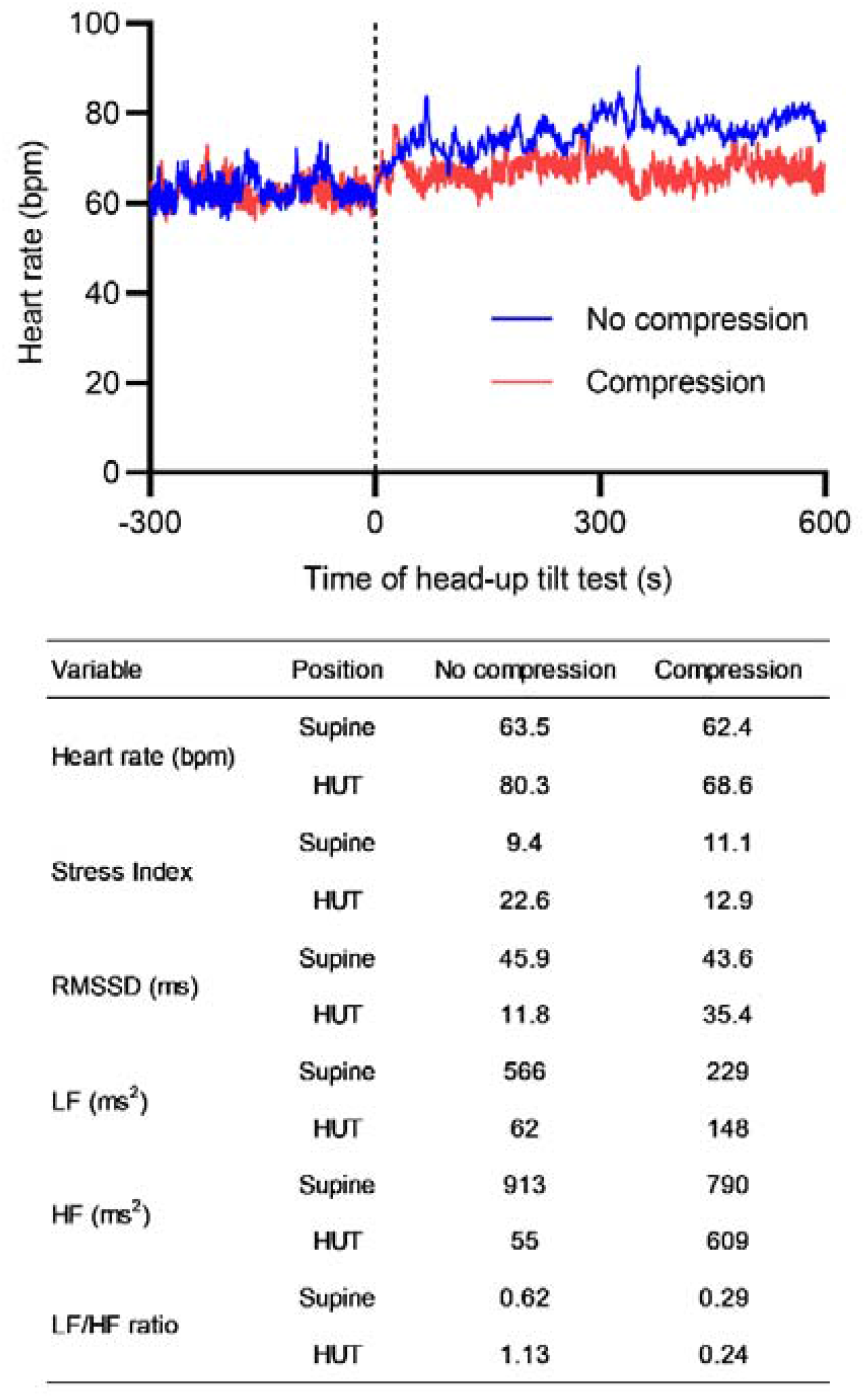
Spectrum of heart rate responses to head-up tilt with and without compression. Representative heart rate changes during the head-up tilt test with (red line) and without (blue line) compression in a female participant. The vertical dashed line indicates the start of the postural change. The table below the figure represents the measurement values of heart rate and heart rate variability parameters. RMSSD, root mean square of successive differences between adjacent R-R intervals; LF, low-frequency component of heart rate variability; HF, high-frequency component of heart rate variability

**Figure 4.**
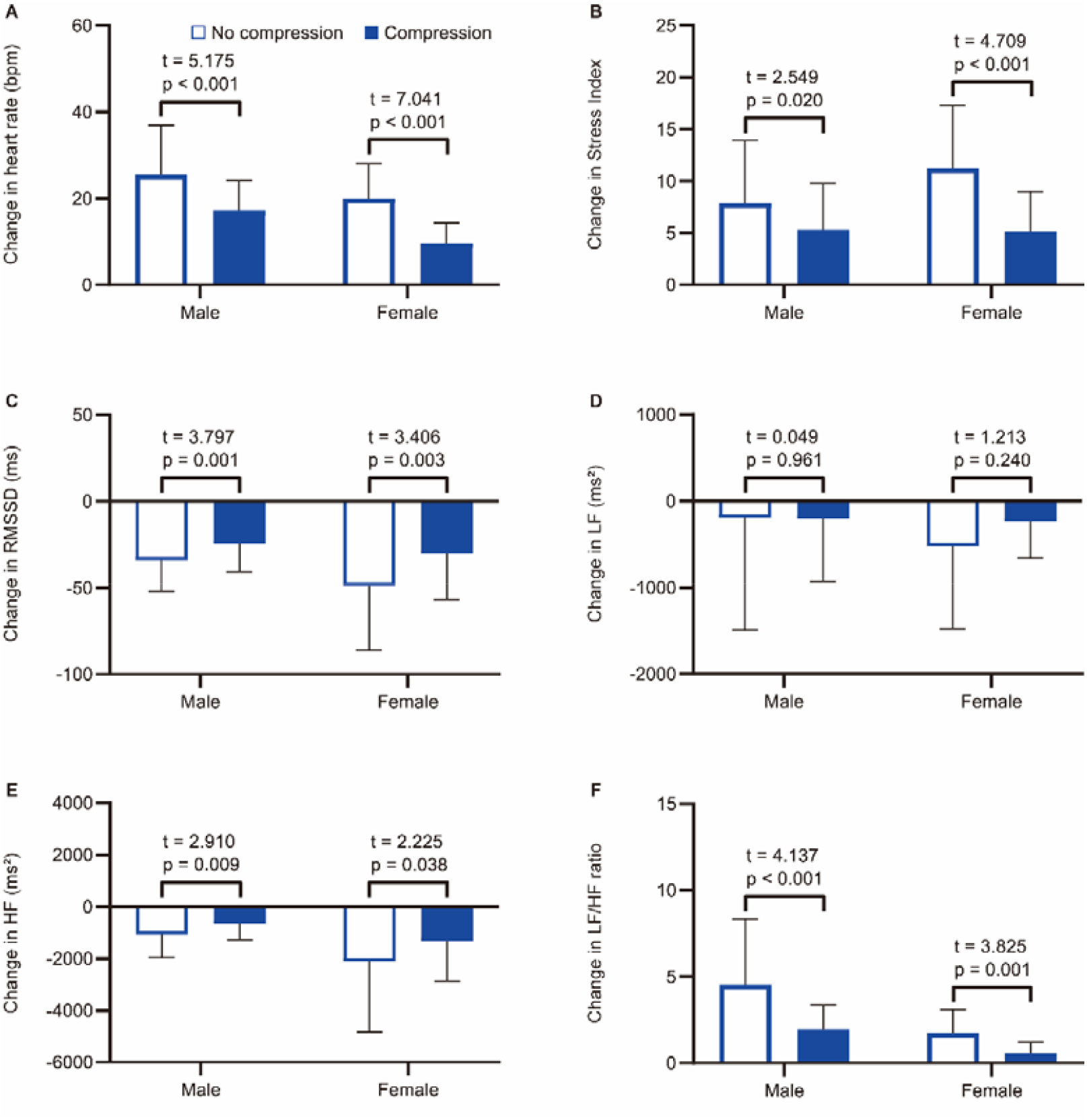
Subgroup analysis by sex for comparisons of the tilt-induced changes in heart rate and heart rate variability parameters with and without compression Changes in (A) heart rate, (B) Stress Index, (C) RMSSD, (D) LF, (E) HF, and (F) LF/HF ratio from supine to head-up tilt. White and blue bars represent the mean values in the no-compression condition and those in the compression condition, respectively. RMSSD, root mean square of successive differences between adjacent R-R intervals; LF, low-frequency component of heart rate variability; HF, high-frequency component of heart rate variability

**Figure 5.**
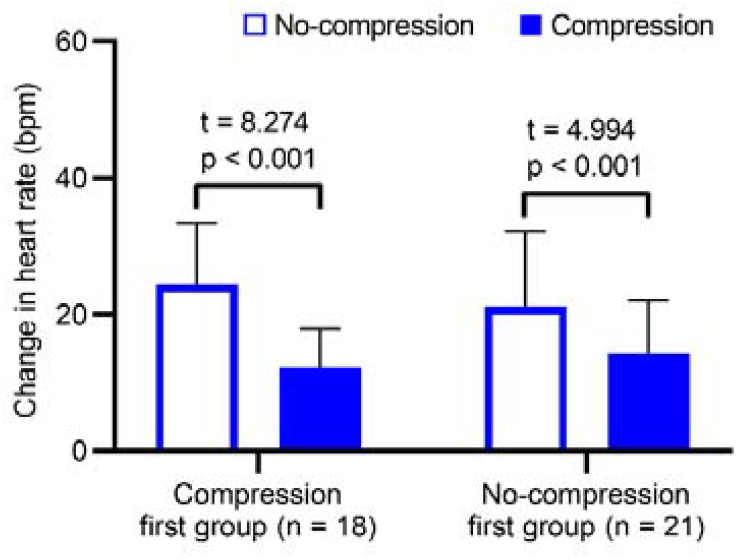
Comparison of heart rate changes during the head-up tilt test between the conditions in the compression-first and no-compression-first groups White and blue bars indicate the mean values of heart rate changes during the head-up tilt test in the no-compression and compression conditions, respectively. Error bars indicate the standard deviation.

Regarding blood pressure variables, there was a statistical main effect of position on systolic [F(1,38) = 55.5, p < 0.001] and diastolic [F(1,38) = 52.5, p < 0.001] blood pressure. For systolic blood pressure, the main effects of the condition [F(1,38) = 13.9, p < 0.001] and the position × condition interaction [F(1,38) = 4.9, p = 0.033] were also significant (Figure 2B), indicating that the reduction in systolic blood pressure was significantly smaller in the compression condition than that in the no-compression condition. In contrast, the main effects of condition [F(1,38) = 0.984, p = 0.328] and the position × condition interaction [F(1,38) = 1.1, p = 0.291] for diastolic blood pressure were not significant (Figure 2C).

### Heart rate variability parameters

The two-way repeated-measures ANOVA yielded significant main effects for position [F(1,38) = 106.4, p < 0.001] and condition [F(1,38) = 9.4, p = 0.004] on the Stress Index, as well as a significant interaction between position and condition [F(1,38) = 25.7, p < 0.001] (Figure 2D). Regarding RMSSD, there was a significant main effect for position [F(1,38) = 77.9, p < 0.001] and a significant interaction between position and condition [F(1,38) = 21.0, p < 0.001]; however, the main effect of condition was not significant [F(1,38) = 1.6, p = 0.213] (Figure 2E). In the supine condition, there were no significant differences between the conditions for both Stress Index (t = 1.076, p = 0.578) and RMSSD (t = 2.300, p = 0.054). During the HUT position, a significantly lower Stress Index (t = 6.091, p < 0.001) and a significantly higher RMSSD (t = 4.176, p < 0.001) were observed in the compression condition than in the no-compression condition. These results indicate that the increase in Stress Index and the decrease in RMSSD during the HUT test were significantly smaller in the compression condition than in the no-compression condition. Subgroup analyses indicated that the increase in Stress Index (Figure 4B) and the decrease in RMSSD (Figure 4C) during the test were significantly smaller in the compression condition than those in the no-compression condition, regardless of sex (p < 0.05).

Concerning the frequency-domain parameters of heart rate variability, the main effect of position on LF was significant [F(1,38) = 6.4, p = 0.016], whereas the main effects of condition [F(1,38) = 0.7, p = 0.402] and the interaction between position and condition [F(1,38) = 0.6, p = 0.439] were not significant (Figure 2F). In the subgroup analysis, the orthostatic change in LF was not statistically significantly different between the conditions, regardless of sex (p > 0.05) (Figure 4D). For HF, the main effects of position [F(1,38) = 26.3, p < 0.001] and the position × condition interaction [F(1,38) = 9.6, p = 0.004] were significant, although there was no significant main effect of condition [F(1,38) = 92.2, p = 0.148] (Figure 2G). A significantly smaller HF in the compression condition than that in the no compression condition was observed during the supine position (t = 3.166, p = 0.006), while there was no significant difference in HF between the conditions during the HUT position (t = 1.225, p = 0.456). Moreover, the results of two-way repeated-measures ANOVA showed a significant main effect of position [F(1,38) = 40.7, p < 0.001], condition [F(1,38) = 25.9, p < 0.001], and the interaction between position and condition [F(1,38) = 27.1, p < 0.001] for LF/HF ratio (Figure 2H). The LF/HF ratio in the supine position was not significantly different between the conditions (t = 0.046, p = 0.999), while that of the HUT position was significantly lower in the compression condition than that in the no-compression condition (t = 7.317, p < 0.001). Subgroup analyses revealed that the decrease in HF (Figure 4E) and the increase in LF/HF ratio (Figure 4F) during the HUT test were significantly smaller in the compression condition than those in the no-compression condition (p < 0.05), regardless of sex.

### Heart rate variability parameters associated with the reduction in heart rate during the HUT position with abdominal and lower-extremity compression

The correlations between heart rate reduction with compression and changes in heart rate variability parameters with compression are shown in Figure 6. There was a significant correlation between a greater reduction in heart rate with compression and a larger decrease in Stress Index (r = 0.552, 95% confidence interval [CI] = 0.286 to 0.739, p < 0.001; Figure 6A). Additionally, an increase in RMSSD with compression correlated with a larger heart rate reduction (r = −0.485, 95% CI = −0.695 to −0.200, p = 0.002; Figure 6B). However, no significant correlations were found between changes in the HF component (r = −0.306, 95% CI = −0.567 to 0.010, p = 0.058; Figure 6C) and the LF/HF ratio (r = 0.115, 95% CI = −0.208 to 0.416, p = 0.486; Figure 6D) with compression. The associations between the heart rate reduction with compression and potentially confounding variables are presented in Table 2. The greater reduction in heart rate measured during the HUT position with compression was also significantly associated with the female sex (mean difference = −4.4, 95% CI = −7.7 to −1.1, p = 0.010) but not with other variables (p > 0.05). Therefore, the changes in Stress Index and RMSSD with compression and sex were entered in the multiple regression analysis as the independent variables. The results of the multiple regression analysis are shown in Table 3. The analysis (adjusted R^2^ = 0.335, p < 0.001) revealed that the decrease in Stress Index with compression was the only significant independent variable for heart rate reduction with compression (coefficient = 0.411, p = 0.025).

**Table 2.**
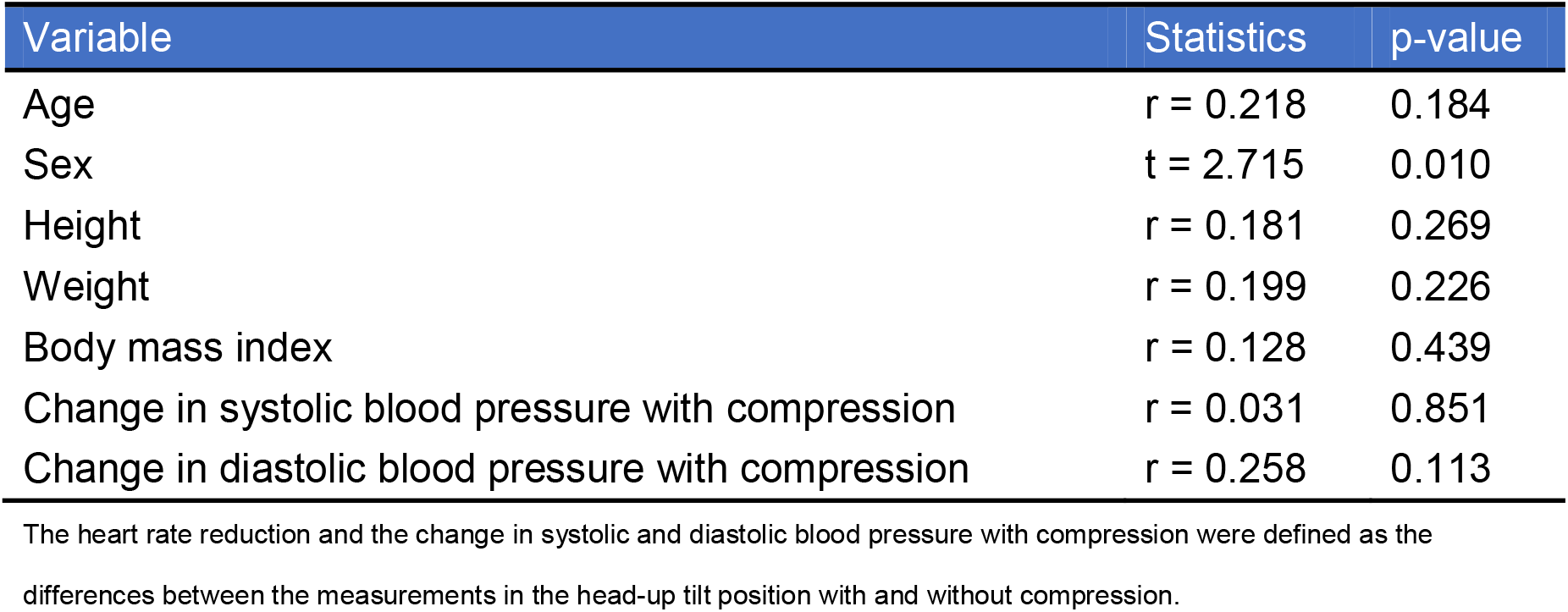
Associations between heart rate reduction with compression and potential confounding variables.

| Variable | Statistics | p-value |
| --- | --- | --- |
| Age | $r = 0.218$ | 0.184 |
| Sex | $t = 2.715$ | 0.010 |
| Height | $r = 0.181$ | 0.269 |
| Weight | $r = 0.199$ | 0.226 |
| Body mass index | $r = 0.128$ | 0.439 |
| Change in systolic blood pressure with compression | $r = 0.031$ | 0.851 |
| Change in diastolic blood pressure with compression | $r = 0.258$ | 0.113 |

The heart rate reduction and the change in systolic and diastolic blood pressure with compression were defined as the differences between the measurements in the head-up tilt position with and without compression.

**Table 3.** Multiple regression analysis to determine the heart rate reduction with compression.

| Variable | Coefficient | SE | t-value | p-value |
| --- | --- | --- | --- | --- |
| Decrease in Stress Index with compression | 0.411 | 0.175 | 2.343 | 0.025 |
| Increase in RMSSD with compression | -0.096 | 0.103 | 0.931 | 0.358 |
| Sex (male = 0, female = 1) | -2.527 | 1.555 | 1.625 | 0.113 |
| Constant | -5.119 | 1.131 | 4.526 | < 0.001 |

$F(3, 35) = 7.4$ , $R^2 = 0.388$ , Adjusted $R^2 = 0.335$ , $p < 0.001$
The heart rate reduction, the decrease in Stress Index, and the increase in RMSSD with compression were defined as the differences between the measurements in the head-up tilt position with and without compression.
SE, Standard error; RMSSD, root mean square of successive differences between adjacent R-R intervals

**Figure 6.**
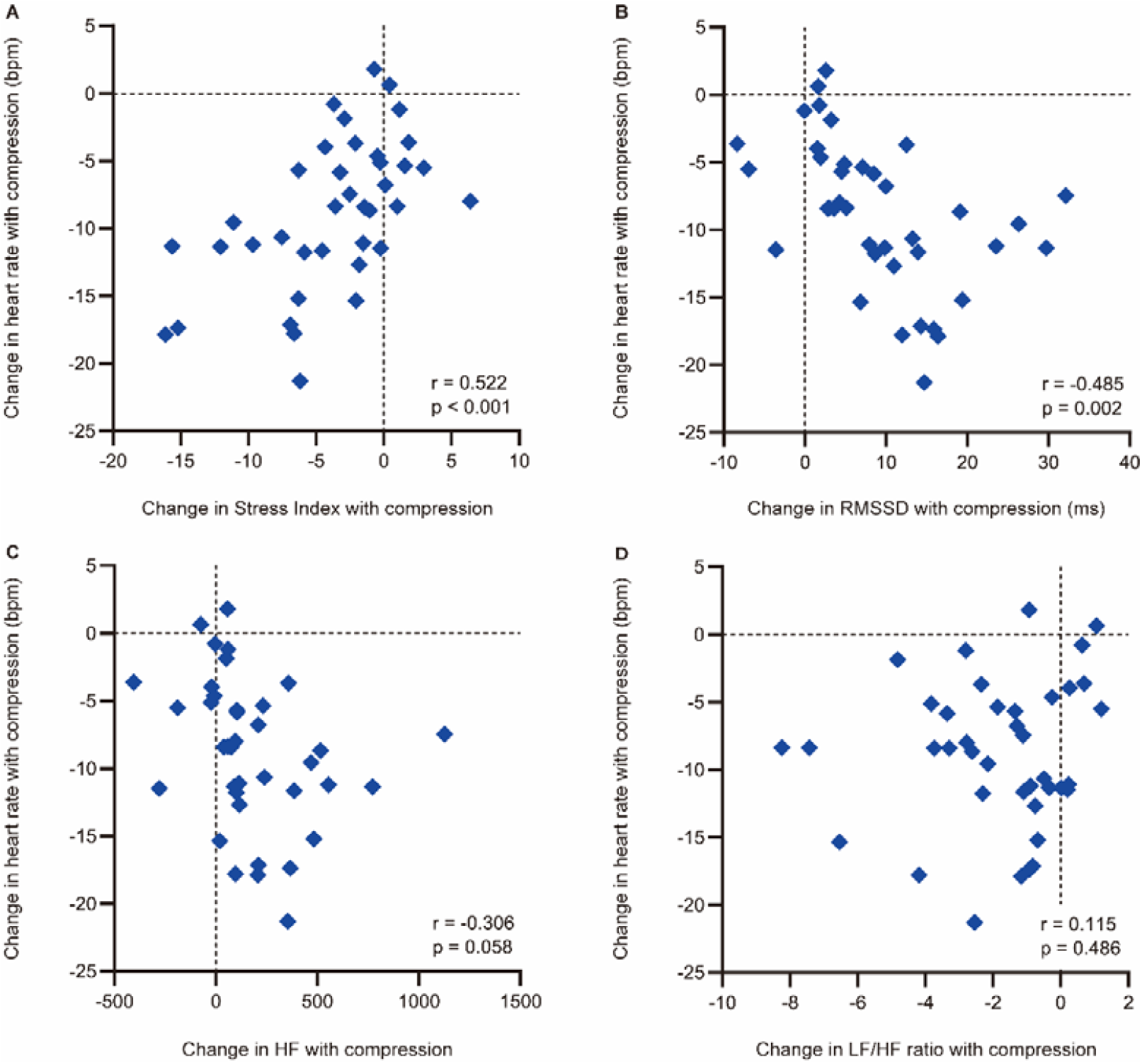
Correlations between the heart rate reduction with compression and the compression-induced changes in heart rate variability parameters with a significant difference with and without compression Correlations between the heart rate reduction with compression and the compression-induced changes in (A) Stress Index, (B) RMSSD, (C) HF, and (D) LF/HF ratio. The heart rate reduction with compression and the compression-induced changes in heart rate variability parameters were defined as the differences between the measurements in the head-up tilt position with and without compression. RMSSD, root mean square of successive differences between adjacent R-R intervals; HF, high-frequency component of heart rate variability

## DISCUSSION

Consistent with previous studies conducted on individuals with POTS (Heyer, 2014; Bourne et al., 2021), our present study found that abdominal and lower-extremity compression effectively reduced the magnitude of heart rate elevation and attenuated the decline in systolic blood pressure during the HUT test. To the best of our knowledge, this study is the first to compare heart rate variability responses to HUT with and without abdominal and lower-extremity compression in healthy individuals. We found that the orthostatic increases in the Stress Index and LF/HF ratio and the orthostatic decreases in RMSSD and HF were smaller in the compression condition than those in the no-compression condition. The reductions in heart rate and alternations in heart rate variability responses to the HUT test with compression were consistent across participants regardless of their sex. Abdominal and lower-extremity compression redistributed blood pooled blood, reducing sympathetic activation and vagal withdrawal, thereby attenuating the heart rate increase during the transition from supine to standing positions.

A crossover design allows a participant’s response to one treatment to be contrasted with the same participant’s response to another. Removing participant variation in this way makes crossover trials potentially more efficient than similar-sized, parallel-group trials in which each participant is exposed to only one treatment (Sibbald and Roberts, 1998). In addition, a rest period of at least 5 min is considered sufficient to establish a stable baseline (Cheshire and Goldstein, 2019; Finucane et al., 2019). Therefore, the HUT tests with and without compression were performed randomly, with a 10-min washout period between each test. These procedures may reduce order bias and carryover effects, which resulted in no significant differences in heart rates and heart rate variability in the supine position between the conditions. We used the inflatable abdominal band with a pressure of 35–45 mmHg and the medical compression stockings with a pressure of approximately 35–45 mmHg at the ankles. A study of healthy adults reported that low-pressure sports compression tights with a pressure of 15.2 ± 7.2 mmHg at the calf and 8.5 ± 1.5 mmHg at the thigh attenuated the increase in heart rate during the HUT test (Lee et al., 2018). Studies of individuals with POTS also demonstrated that abdominal and lower-extremity compression with a pressure of 20 to 40 mmHg reduced orthostatic tachycardia during the HUT test (Heyer, 2014; Bourne et al., 2021). Given the results of these previous studies, the compression pressure used in this study might be sufficient to reduce the orthostatic increase in heart rate.

As the increase in the Stress Index from a supine to a standing position is thought to be a sensitive marker for orthostatic sympathetic activation (Baevsky and Chernikova, 2017; Ali et al., 2021), the results of multiple regression analysis suggest that the decrease in cardiac sympathetic activity is associated with the heart rate reduction in the HUT position with compression. This may help provide insights into the mechanisms underlying the improvements in an excessive orthostatic heart rate increase with compression. Excessive central sympathetic activation is one of the possible mechanisms leading to POTS, which is called hyperadrenergic POTS (Lambert and Lambert, 2014; Sheldon et al., 2015; Arnold et al., 2018; Bryarly et al., 2019; Vernino et al., 2021). Central sympatholytic agents, such as clonidine and methyldopa, improve orthostatic tachycardia in individuals with hyperadrenergic POTS (Sheldon et al., 2015; Vernino et al., 2021). However, non-pharmacological therapies are often recommended as a first-line treatment for POTS (Sheldon et al., 2015; Fu and Levine, 2018; Bryarly et al., 2019; Vernino et al., 2021). Considering our results of multiple regression analysis, abdominal and lower-extremity compression may be a beneficial treatment for individuals with hyperadrenergic POTS. Further studies are needed to examine whether abdominal and lower-extremity compression can reduce these individuals’ excessive central sympathetic activity and orthostatic tachycardia.

This study had some limitations. First, we evaluated only healthy adults to demonstrate normal compression-induced changes in heart rate variability responses to HUT. Further studies of individuals with POTS are warranted to confirm the robustness of our findings. Second, blinding participants to the compression condition was not feasible in our study because of the visible and tangible nature of the inflatable abdominal band and medical compression stockings. Even sham garments without compression would have felt distinct and not allowed for effective blinding. Thus, any placebo effects might have contributed to the results. However, this appears to be consistent with the actual situation of the use of abdominal and lower-extremity compression. Finally, the tests were performed in the evening. Previous research has demonstrated that the orthostatic increase in heart rate tends to be greater in the morning compared to that in the afternoon (Brewster et al., 2012). Given that a higher heart rate increase from supine to HUT in the no-compression condition is associated with a greater heart rate reduction in the HUT position with compression (Bourne et al., 2021), this study may underestimate the effects of compression on heart rate and heart rate variability responses to HUT.

In conclusion, this study demonstrated that the heart rate and heart rate variability responses to HUT were smaller in the compression condition than those in the no-compression condition. These results suggest that abdominal and lower-extremity compression attenuates orthostatic cardiac sympathetic activation and orthostatic decrease in vagal activity in healthy adults. In addition, our results also indicate that the decreased cardiac sympathetic activity may be associated with the attenuation of the orthostatic heart rate increase with compression, which will contribute to understanding the mechanisms underlying the improvements of an excessive orthostatic heart rate increase with compression. The findings of this study may have important implications for the application of abdominal and lower-extremity compression to prevent the development of orthostatic tachycardia.

## Author Contributions

KO, MK, AY, and KM contributed to the conception and design of the study. KO, MK, AY, HI, and SK contributed to data acquisition, analysis, and interpretation. KO obtained funding and wrote the first draft of the manuscript. All authors contributed to manuscript revision and read and approved the submitted manuscript.

## Acknowledgments

We would like to thank Editage (www.editage.com) for English language editing.

## Data Availability Statement

The original contributions represented in the study are included in the article/supplementary material; further inquiries can be directed to the corresponding author.

